# Effectiveness of Virtual Reality in Pain Management, Anxiety Control, and Patient Satisfaction in Bronchoscopy: A Systematic Review Protocol

**DOI:** 10.1101/2025.07.07.25331075

**Authors:** Ravi Shankar, Fiona Devi, Xu Qian

## Abstract

Bronchoscopy is a common invasive procedure that can cause significant anxiety, discomfort, and pain in patients. Virtual reality (VR) has emerged as a promising non-pharmacological intervention to alleviate these negative experiences and improve patient outcomes. This systematic review aims to comprehensively synthesize the current evidence on the effectiveness of VR in enhancing pain management, anxiety control, and patient satisfaction during bronchoscopy procedures.

The review will follow the Preferred Reporting Items for Systematic Reviews and Meta-Analyses (PRISMA) guidelines and systematically search multiple electronic databases, including PubMed, Web of Science, Embase, CINAHL, MEDLINE, The Cochrane Library, PsycINFO, and Scopus, from their inception to July 2025. Randomized controlled trials (RCTs) comparing VR interventions to standard care or other non-pharmacological interventions in adult patients undergoing bronchoscopy will be included.

The primary outcomes of interest are pain intensity, anxiety levels, and patient satisfaction, measured using validated scales or questionnaires. Secondary outcomes will include physiological parameters, procedure duration, medication use, and adverse events related to VR interventions. Two reviewers will independently screen studies, extract data, and assess risk of bias using the Cochrane Risk of Bias tool for RCTs and the ROBINS-I tool for non-randomized studies.

If feasible, a meta-analysis will be conducted to quantitatively synthesize the results. Subgroup and sensitivity analyses will be performed to explore potential sources of heterogeneity and assess the robustness of the findings. The Grading of Recommendations, Assessment, Development, and Evaluation (GRADE) approach will be used to evaluate the overall certainty of evidence for each outcome.

This systematic review protocol is guided by established frameworks, including the PICO (Population, Intervention, Comparison, Outcome), PRISMA-P (Preferred Reporting Items for Systematic Review and Meta-Analysis Protocols), and FINER (Feasible, Interesting, Novel, Ethical, and Relevant) criteria. By providing a transparent and rigorous evaluation of the effectiveness of VR in improving patient experience during bronchoscopy, this review will offer valuable insights to inform clinical practice, guide health policy decisions, and identify areas for future research.

## Introduction

### Background and Rationale

Bronchoscopy is a widely used invasive procedure for the diagnosis and treatment of various respiratory diseases, including lung cancer, infections, and airway disorders [1,2]. It involves the insertion of a flexible or rigid tube called a bronchoscope through the nose or mouth into the airways, allowing direct visualization and sampling of the tracheobronchial tree [3]. While generally safe and well-tolerated, bronchoscopy can induce significant anxiety, discomfort, and pain in patients due to the invasive nature of the procedure and the stimulation of cough receptors in the airways [4,5].

Inadequate management of pain and anxiety during bronchoscopy can lead to several adverse consequences. Patients may experience heightened distress, fear, and dissatisfaction with the procedure, which can result in premature termination, suboptimal examination, and avoidance of necessary follow-up bronchoscopies [6,7]. Moreover, excessive patient movement and coughing during the procedure can increase the risk of complications, prolong the procedure duration, and compromise the quality of diagnostic specimens [8].

Current strategies for managing pain and anxiety during bronchoscopy primarily rely on pharmacological interventions, such as sedatives, analgesics, and local anesthetics [9]. While these medications can effectively reduce patient discomfort, they also have potential drawbacks. Sedatives may cause respiratory depression, cardiovascular instability, and prolonged recovery time, necessitating additional monitoring and resources [10]. Local anesthetics can have limited effectiveness in controlling cough reflexes and may cause unpleasant taste and numbness [11]. Furthermore, the use of pharmacological agents increases the overall cost of the procedure and may not be suitable for all patients, particularly those with comorbidities or allergies [12].

Given these limitations, there is a growing interest in exploring non-pharmacological interventions that can complement or replace pharmacological approaches to improve patient experience during bronchoscopy. Non-pharmacological interventions, such as music therapy, relaxation techniques, and distraction methods, have shown promise in reducing pain and anxiety in various medical procedures [13,14]. However, their effectiveness in the specific context of bronchoscopy remains unclear.

Virtual reality (VR) has emerged as a particularly promising non-pharmacological intervention for managing pain and anxiety in healthcare settings [15]. VR refers to a computer-generated simulation of a three-dimensional environment that can be explored and interacted with by a person using specialized equipment, such as a head-mounted display and motion sensors [16]. By immersing patients in an engaging and interactive virtual environment, VR can distract them from the negative stimuli associated with medical procedures, thereby reducing pain perception and anxiety levels [17].

Several studies have investigated the use of VR in various medical procedures, such as burn wound care [18], dental treatment [19], and cancer chemotherapy [20], with encouraging results. However, the evidence regarding the effectiveness of VR in the specific context of bronchoscopy is limited and inconsistent. Some studies have reported significant reductions in pain and anxiety with VR interventions during bronchoscopy [21,22], while others have found no significant benefits compared to standard care or other non-pharmacological interventions [23,24].

To date, no systematic review has comprehensively synthesized the available evidence on the effectiveness of VR in bronchoscopy. Previous reviews have either focused on VR in other medical procedures [25] or included only a small number of studies specific to bronchoscopy [26]. A rigorous and up-to-date systematic review is needed to critically appraise the current literature, provide a quantitative synthesis of the results, and identify gaps in knowledge to guide future research and clinical practice.

### Objectives

The primary objective of this systematic review is to evaluate the effectiveness of virtual reality interventions in reducing pain intensity, anxiety levels, and improving patient satisfaction during bronchoscopy procedures in adult patients compared to standard care or other non-pharmacological interventions.

The specific research questions are:

1. Do VR interventions reduce pain intensity during bronchoscopy compared to standard care or other non-pharmacological interventions?
2. Do VR interventions reduce anxiety levels during bronchoscopy compared to standard care or other non-pharmacological interventions?
3. Do VR interventions improve patient satisfaction with bronchoscopy compared to standard care or other non-pharmacological interventions?
4. Are there any adverse events associated with the use of VR interventions during bronchoscopy?

The secondary objectives are to explore the effects of VR interventions on physiological parameters, procedure duration, and medication use during bronchoscopy, as well as to identify potential moderators of the effectiveness of VR interventions, such as the type of VR system, patient characteristics, and procedural factors.

## Methods

### Protocol and Registration

This systematic review protocol follows the Preferred Reporting Items for Systematic Review and Meta-Analysis Protocols (PRISMA-P) guidelines [27] to ensure transparency, reproducibility, and quality. The protocol has been registered in the International Prospective Register of Systematic Reviews (PROSPERO) database to avoid duplication and promote accountability (registration number: CRD42025636320).

### Eligibility Criteria

The eligibility criteria for studies to be included in this systematic review are defined using the PICO (Population, Intervention, Comparison, Outcome) framework [28].

### Population

The population of interest is adult patients (aged 18 years or older) undergoing bronchoscopy for diagnostic or therapeutic purposes. Studies involving pediatric patients or patients undergoing other types of endoscopic procedures will be excluded.

### Intervention

The intervention of interest is virtual reality-based interventions used for pain management, anxiety reduction, or patient distraction during bronchoscopy. VR interventions can be either immersive (using head-mounted displays) or non-immersive (using desktop or projection systems) and may involve various types of virtual environments, such as natural landscapes, relaxation scenes, or interactive games.

Studies that combine VR interventions with pharmacological agents will be included if the co-interventions are equally applied in both the VR and control groups. Studies that do not provide sufficient details about the VR intervention will be excluded.

### Comparison

The comparison groups of interest include:

1. Standard care alone (e.g., pharmacological interventions, such as sedatives or analgesics)
2. Other non-pharmacological interventions (e.g., music therapy, relaxation techniques, distraction methods)
3. Sham VR interventions (e.g., non-immersive video viewing, static images)

Studies that do not have a control group or do not clearly define the comparison intervention will be excluded.

### Outcomes

#### The primary outcomes of interest are

1. Pain intensity, measured by validated self-report scales, such as the Visual Analog Scale (VAS), Numeric Rating Scale (NRS), or McGill Pain Questionnaire (MPQ)
2. Anxiety levels, measured by validated self-report scales, such as the State-Trait Anxiety Inventory (STAI), Hospital Anxiety and Depression Scale (HADS), or Visual Analog Scale for Anxiety (VAS-A)
3. Patient satisfaction, measured by validated questionnaires, such as the Patient Satisfaction Questionnaire (PSQ) or Likert-type scales

#### The secondary outcomes of interest include

1. Physiological parameters, such as heart rate, blood pressure, respiratory rate, and oxygen saturation
2. Procedure duration, measured in minutes or seconds
3. Medication use, including the type and dosage of sedatives, analgesics, or local anesthetics
4. Adverse events related to the VR intervention, such as motion sickness, eye strain, or disorientation

### Study Design

Randomized controlled trials (RCTs) comparing VR interventions to standard care, other non-pharmacological interventions, or sham VR interventions will be included. Cluster RCTs and crossover trials will also be eligible if they have appropriate washout periods and account for potential carryover effects.

Non-randomized studies, such as cohort studies, case-control studies, and before-after studies, will be considered for inclusion if they have a clearly defined control group and adjust for potential confounders. Case reports, case series, qualitative studies, and reviews will be excluded.

### Information Sources and Search Strategy

A comprehensive literature search will be conducted in the following electronic databases from their inception to July 2025: PubMed (including MEDLINE), Web of Science, Embase, CINAHL, PsycINFO, Scopus, and the Cochrane Library (including the Cochrane Central Register of Controlled Trials and the Cochrane Database of Systematic Reviews).

The search strategy will be developed in collaboration with a medical librarian and will include a combination of keywords and controlled vocabulary terms (e.g., MeSH terms) related to bronchoscopy, virtual reality, pain, anxiety, and patient satisfaction. The following search string template will be adapted for each database:

((“bronchoscopy” OR “bronchoscopic” OR “airway examination” OR “lung examination”) AND (“virtual reality” OR “VR” OR “head-mounted display” OR “immersive virtual environment” OR “3D environment”) AND (“pain” OR “analgesia” OR “discomfort” OR “anxiety” OR “distress” OR “satisfaction”) AND (“randomized controlled trial” OR “RCT” OR “randomized” OR “randomly”))

In addition to the electronic database search, we will manually search the reference lists of included studies and relevant systematic reviews to identify any additional eligible studies. We will also search for ongoing or unpublished trials in clinical trial registries, such as ClinicalTrials.gov and the World Health Organization International Clinical Trials Registry Platform.

No language or publication date restrictions will be applied to the search. Non-English studies will be translated using professional translation services. Studies published in peer-reviewed journals, conference proceedings, and dissertations will be considered for inclusion.

### Study Selection

The study selection process will consist of two stages: title and abstract screening and full-text review. In the first stage, two reviewers will independently screen the titles and abstracts of all retrieved records against the predefined eligibility criteria. Studies that clearly do not meet the inclusion criteria will be excluded. Any disagreements between the reviewers will be resolved through discussion or by consulting a third reviewer.

In the second stage, the full-text articles of the remaining studies will be obtained and assessed for eligibility by two independent reviewers. The reasons for exclusion will be recorded and reported in the final review. Any discrepancies between the reviewers will be resolved through discussion or by involving a third reviewer.

The study selection process will be documented using a PRISMA flow diagram [29], which will outline the number of studies identified, screened, eligible, and included in the review. The Covidence systematic review software [30] will be used to manage the screening and data extraction process.

### Data Extraction

A standardized data extraction form will be developed and piloted on a sample of included studies to ensure its completeness and reliability. Two reviewers will independently extract data from the included studies, and any discrepancies will be resolved through discussion or by consulting a third reviewer.

The extracted data will encompass several key domains to ensure a comprehensive synthesis of the included studies. These will include: (1) study characteristics, such as first author, year of publication, country, sample size, study design, and funding source; (2)participant characteristics, including age, gender, ethnicity, indication for bronchoscopy, and relevant comorbidities; (3) intervention details, such as the type of VR system used, duration and frequency of VR sessions, type of virtual environment, and any co-interventions; (4) comparison details, including the nature of standard care or other non-pharmacological interventions and their session characteristics; (5) outcome measures, covering variables such as pain intensity, anxiety levels, patient satisfaction, physiological parameters, procedure duration, medication use, and adverse events; (6) results, including statistical data such as means and standard deviations, effect sizes, p-values, and confidence intervals; and (7) methodological quality, assessing aspects such as risk of bias, allocation concealment, and blinding.

If any essential data are missing or unclear, we will contact the study authors for clarification or additional information. If the authors do not respond after two attempts, the study will be excluded from the analysis.

### Risk of Bias Assessment

The risk of bias in the included studies will be assessed using appropriate tools depending on the study design. For RCTs, the Cochrane Risk of Bias tool (RoB 2) [31] will be used, which evaluates six domains of bias: randomization process, deviations from intended interventions, missing outcome data, measurement of the outcome, selection of the reported result, and overall bias. Each domain will be judged as low risk, high risk, or some concerns.

For non-randomized studies, the Risk Of Bias In Non-randomized Studies of Interventions (ROBINS-I) tool [32] will be employed. ROBINS-I assesses seven domains of bias: confounding, selection of participants, classification of interventions, deviations from intended interventions, missing data, measurement of outcomes, and selection of reported results. Each domain is judged as low, moderate, serious, or critical risk of bias.

Two reviewers will independently assess the risk of bias in each included study, and any disagreements will be resolved through discussion or by involving a third reviewer. The results of the risk of bias assessment will be presented in a risk of bias summary table and figure, which will highlight the overall quality of the evidence.

### Data Synthesis

If the included studies are sufficiently homogeneous in terms of participants, interventions, comparisons, and outcomes, a meta-analysis will be conducted to quantitatively synthesize the results. Continuous outcomes (e.g., pain intensity, anxiety levels) will be analyzed using mean differences (MD) or standardized mean differences (SMD) with 95% confidence intervals (CI), depending on whether the studies used the same or different measurement scales. Dichotomous outcomes (e.g., adverse events) will be analyzed using risk ratios (RR) with 95% CI.

Heterogeneity among the studies will be assessed using the I^2^ statistic and the Chi^2^ test. An I^2^ value greater than 50% or a p-value less than 0.10 in the Chi^2^ test will be considered indicative of substantial heterogeneity [33]. If significant heterogeneity is detected, a random-effects model will be used for the meta-analysis, as it accounts for both within-study and between-study variability. If there is no significant heterogeneity, a fixed-effect model will be employed.

If the included studies are too heterogeneous to be combined in a meta-analysis, a narrative synthesis will be conducted. The narrative synthesis will focus on describing the direction, magnitude, and consistency of the effects across the studies, as well as exploring the potential sources of heterogeneity.

### Subgroup and Sensitivity Analyses

If sufficient data are available, subgroup analyses will be performed to investigate the potential moderators of the effectiveness of VR interventions. The planned subgroup analyses include:

1. Type of VR system (e.g., immersive vs. non-immersive, head-mounted display vs. desktop)
2. Type of virtual environment (e.g., natural scenes vs. interactive games)
3. Duration and frequency of VR sessions
4. Patient characteristics (e.g., age, gender, indication for bronchoscopy)
5. Procedural factors (e.g., type of bronchoscope, use of sedation)

Sensitivity analyses will be conducted to assess the robustness of the findings by excluding studies with high risk of bias, small sample sizes, or outlying results. If the removal of these studies significantly alters the overall effect estimates, the results will be interpreted with caution.

### Meta-Bias Assessment

To assess the potential for publication bias, funnel plots will be generated if there are at least ten studies included in the meta-analysis. Asymmetry in the funnel plot may indicate the presence of publication bias, which will be further examined using Egger’s test [34]. If publication bias is detected, the trim-and-fill method [35] will be applied to estimate the impact of missing studies on the pooled effect size.

Other sources of bias, such as selective outcome reporting and language bias, will also be considered. We will compare the outcomes reported in the published articles with those listed in the study protocols or trial registries to identify any discrepancies. The potential for language bias will be assessed by examining the proportion of non-English studies included in the review.

### Confidence in Cumulative Evidence

The strength of the body of evidence for each outcome will be assessed using the Grading of Recommendations, Assessment, Development, and Evaluation (GRADE) approach [36]. The GRADE system considers five domains: risk of bias, inconsistency, indirectness, imprecision, and publication bias. Based on these domains, the quality of evidence will be rated as high, moderate, low, or very low.

Two reviewers will independently assess the quality of evidence for each outcome, and any disagreements will be resolved through discussion or by consulting a third reviewer. The results of the GRADE assessment will be presented in a summary of findings table, which will provide a concise overview of the key findings and the confidence in the estimates.

## Discussion

This systematic review protocol presents a comprehensive and rigorous plan to synthesize the available evidence on the effectiveness of virtual reality interventions in reducing pain, anxiety, and improving patient satisfaction during bronchoscopy procedures. By adhering to established guidelines and frameworks, such as PRISMA-P, PICO, and FINER, this protocol aims to ensure the transparency, reproducibility, and quality of the review process.

One of the strengths of this protocol is the extensive literature search strategy, which encompasses multiple electronic databases, manual searching of reference lists, and exploration of grey literature sources. This approach minimizes the risk of missing relevant studies and allows for a comprehensive evaluation of the current state of evidence. Additionally, the involvement of a medical librarian in developing the search strategy enhances its sensitivity and specificity.

Another strength is the use of standardized tools and methods for data extraction, risk of bias assessment, and quality of evidence appraisal. The Cochrane RoB 2 tool and the ROBINS-I tool are well-established and validated instruments for assessing the risk of bias in randomized and non-randomized studies, respectively. The GRADE approach provides a systematic and transparent framework for evaluating the confidence in the cumulative evidence. The use of these tools ensures that the review findings are based on the best available evidence and are critically appraised for their methodological quality.

However, there are also potential limitations and challenges that should be acknowledged. First, the anticipated heterogeneity among the included studies in terms of VR interventions, comparators, and outcome measures may pose difficulties in pooling the results and drawing definitive conclusions. To address this issue, we plan to conduct subgroup and sensitivity analyses to explore the potential sources of heterogeneity and assess the robustness of the findings. If the heterogeneity is substantial, a narrative synthesis will be performed instead of a meta-analysis.

Second, the rapid advancement of VR technology and its increasing application in healthcare settings may result in a large number of studies being published in the near future. As this systematic review includes studies published up to July 2025, it may not capture the most recent developments in the field. To mitigate this limitation, we plan to update the review periodically to incorporate new evidence and ensure its relevance to current practice.

Third, the focus of this review is on the short-term effects of VR interventions during bronchoscopy procedures. The long-term impact of VR on patient outcomes, such as post-procedure pain, anxiety, or quality of life, is not addressed. Future studies and reviews may need to investigate the potential long-term benefits or harms of VR interventions in this context.

Despite these limitations, this systematic review has significant implications for clinical practice, research, and health policy. If VR interventions are found to be effective in reducing pain, anxiety, and improving patient satisfaction during bronchoscopy, they could be incorporated into standard care protocols as an adjunctive or alternative approach to pharmacological interventions. This could lead to a reduction in medication use, shorter procedure times, and improved patient experiences.

Furthermore, the identification of specific VR features or patient characteristics that moderate the effectiveness of VR interventions could guide the development and implementation of tailored VR programs for different patient populations or settings. This information could also inform the design of future RCTs to test the efficacy of optimized VR interventions against current best practices.

From a health policy perspective, the evidence generated by this systematic review could support the allocation of resources and funding for the integration of VR technology into healthcare systems. If VR interventions are cost-effective and improve patient outcomes, they could be prioritized as a valuable investment in patient care.

In conclusion, this systematic review protocol outlines a rigorous and comprehensive approach to synthesizing the evidence on the effectiveness of virtual reality interventions in bronchoscopy procedures. The findings of this review have the potential to advance our understanding of the role of VR in managing pain, anxiety, and patient satisfaction during invasive medical procedures. By providing a solid foundation for evidence-based practice and decision-making, this review can ultimately contribute to improving the quality and patient-centeredness of bronchoscopy care.

## Data Availability

All data produced in the present study are available upon reasonable request to the corresponding author.

---

**Appendix 1.** Data Extraction Form.

| Category | Items |
| --- | --- |
| **1. Study ID** | First author<br>Year of publication<br>Country |
| **2. Methods** | Study design<br>Sample size<br>Setting<br>Study duration |
| **3. Participants** | Age (mean, SD, range)<br>Gender (% male)<br>Inclusion criteria<br>Exclusion criteria |
| **4. Intervention** | Type of VR system<br>Type of virtual environment<br>Duration and frequency of VR sessions<br>Co-interventions |
| **5. Comparison** | Type of control intervention<br>Duration and frequency of control sessions |
| **6. Outcomes** | **Primary outcomes** (e.g., pain intensity, anxiety levels, patient satisfaction)<br>– Measurement tools<br>– Time points<br>**Secondary outcomes** (e.g., physiological parameters, procedure duration, medication use, adverse events)<br>– Measurement tools<br>– Time points |
| **7. Results** | Baseline characteristics<br>Primary outcome results<br>Secondary outcome results<br>Adverse events |
| **8. Miscellaneous** | Funding source<br>Conflicts of interest<br>Author conclusions<br>Reviewer comments |

## Notes

### Competing Interest Statement

The authors have declared no competing interest.

### Funding Statement

This study did not receive any funding

